# Prognostic Impact of Quantitative CT Disease Extent and Pattern in Hypersensitivity Pneumonitis

**DOI:** 10.1101/2025.10.12.25337853

**Authors:** Evans R. Fernández Pérez, Stephen M. Humphries, David A. Lynch, Teng Moua, Sachin Chaudhary, Traci N. Adams, Ayodeji Adegunsoye, Mary Beth Scholand, Namita Sood, Brian Vestal

## Abstract

**Rationale:** In contrast to visual assessments, quantitative CT using multiple instance learning (MIL)-usual interstitial pneumonia (UIP) classifier and data-driven textural analysis (DTA) provides objective and reproducible measurements of disease pattern and extent, respectively. However, the understanding of their individual or combined relationships with outcomes in hypersensitivity pneumonitis (HP) remains limited.

**Objectives:** To determine whether visual CT patterns of HP or UIP, a MIL-UIP classifier, baseline and changes in DTA over 12 months and DTA at 12 months are associated with progression-free survival (PFS) in HP.

**Methods:** The primary cohort included 134 participants with HP. Associations between CT variables and subsequent lung function change, and PFS were assessed using linear mixed models and Cox hazards models. Results were validated in a combined dataset from three independent populations (N=216).

**Results:** Visual patterns of HP or UIP were not associated with PFS. A baseline MIL-UIP classification and DTA were significantly associated with 24-month PFS. However, a MIL-UIP classifier did not significantly predict PFS after adjusting for extent of fibrosis measured by DTA. Over 12 months, a 1-point increase in DTA across and within patients resulted in a significant decline in FVC% and DLCO%. A 12-month change in DTA (HR=1.07, 95%CI 1.01-1.13) and DTA at 12-months (HR=1.03, 95%CI 1.01-1.05) were associated with subsequent 12-month PFS. Similar estimates were observed in the validation cohort.

**Conclusions:** In HP, the objectively measured extent of lung fibrosis is more prognostic than either a UIP classifier or visual CT patterns of HP or UIP. Baseline DTA, DTA at 12 months, and its change over 12 months are associated with decreased PFS.

## BACKGROUND

Hypersensitivity pneumonitis (HP) is a complex, immunologically mediated form of lung disease resulting from repeated inhalational exposure to a wide variety of inciting antigens. A subgroup develops progressive pulmonary fibrosis, which is a leading cause of morbidity and mortality.^1-4^ Recent guidelines have established CT criteria for classifying fibrotic HP (fHP) into three patterns: typical, compatible, and indeterminate.^5^ These patterns provide a qualitative assessment and may suggest the potential behavior of the disease. Notably, a usual interstitial pneumonia (UIP)-like pattern (e.g., “indeterminate for HP”) is associated with a poor prognosis, similar to IPF. Moreover, several studies have suggested that in patients with fHP, a UIP-like fibrotic pattern on HRCT is associated with more rapid disease progression and higher mortality than other fibrotic patterns.^6,7^

While several studies involving diverse populations with ILD have assessed the predictive value of either a UIP-like pattern or isolated HRCT features including the extent fibrosis,^3,4,8-11^ only one study has evaluated the impact of visual UIP in conjunction with the quantitative extent of fibrosis ILD.^12^ However, the majority of HRCT and non-HRCT prognostic studies in HP have, to date, not accounted for the baseline extent of CT fibrosis, which has been shown to be predictive of worse outcomes. Furthermore, there have been no studies investigating the combined effect of a quantitative HRCT UIP pattern and the quantitative extent of fibrosis, or the change in the quantitative extent of fibrosis and its subsequent outcome in well-characterized patients with HP.

Compared to HRCT visual assessments of the extent of fibrosis, quantitative assessments provide a more objective evaluation of the disease’s severity and progression. Our group has developed a machine learning algorithm, data-driven texture analysis (DTA), capable of quantifying the extent of lung fibrosis on HRCT, shown to be an accurate and reproducible measure of disease severity and progression.^13-16^ Also, we have recently demonstrated that multiple instance learning (MIL),^17^ a supervised deep learning technique applied to CT scans, is more sensitive and accurate in classifying a radiologic HRCT UIP patterns than traditional visual assessments. This methodology yields valuable prognostic information that complements quantitative HRCT analysis of fibrosis extent. Nonetheless, the relationships between DTA and MIL-UIP as indicators of fHP severity and predictors of outcomes in fHP remain untested in prospective and independent validation cohorts.

In this study, we aimed to determine whether visual HRCT patterns of HP or UIP, as well as a MIL-UIP classifier, and both baseline and changes in DTA fibrosis scores over 12 months, as well as DTA scores at 12 months, are associated with progression-free survival in a multicenter cohort of HP patients.

## METHODS

### Primary cohort

The primary cohort consisted of a multicenter 24-month prospective observational study conducted from December 2020 to June 2025 across seven centers in the U.S. involving 134 adult patients diagnosed with fHP (NCT04844359),^18^ **Figure 1**. The study was approved by the Mayo Clinic Institutional Review Board (#20-004479) and the local Institutional Review Boards before enrollment at each site.

**Figure 1.**
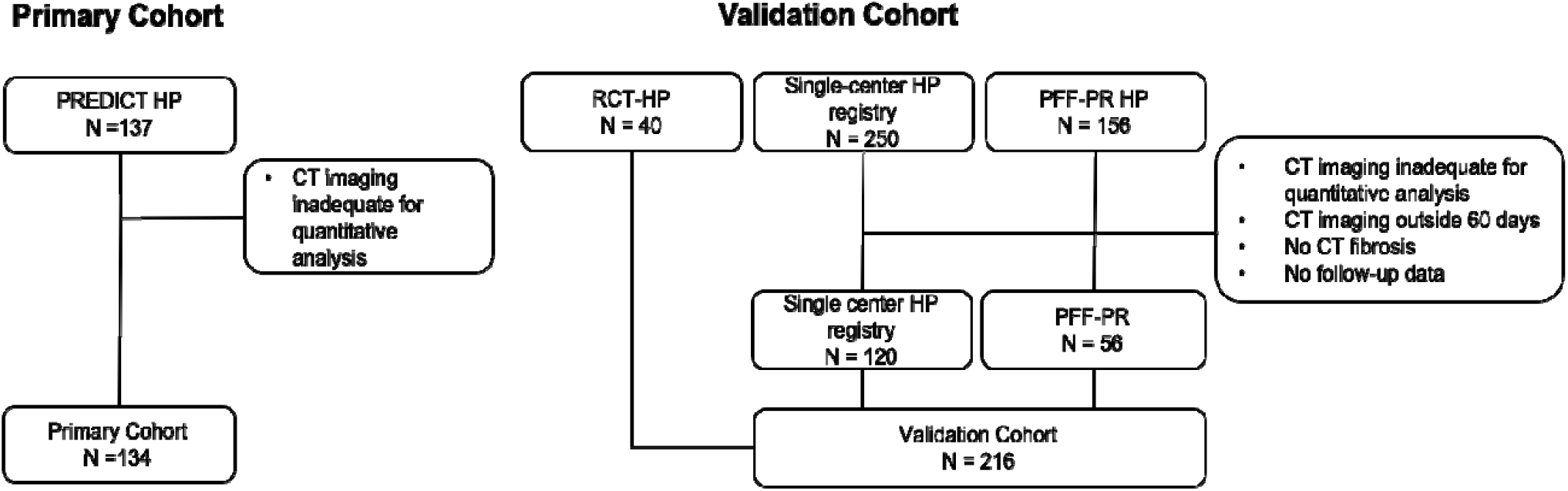
CONSORT (Consolidated Standards of Reporting Trials) diagram for the primary and validation cohorts. CT = computed tomography; HP = hypersensitivity pneumonitis; PFF-PR = Pulmonary foundation patient registry; PREDICT = Prognostic transcriptomic signature in fibrotic hypersensitivity pneumonitis (NCT04844359). RCT = randomized clinical trial (NCT02958917).

For inclusion, patients were required to have a multidisciplinary diagnosis of fHP according to diagnostic guidelines for inclusion. They received standard care for HP, as determined by their treating physicians. At the time of study enrollment and at 12 months, patients underwent clinical assessments, pulmonary function testing, and a chest HRCT chest. All patients provided written informed consent before participating in the study.

### Validation cohort

*Baseline HRCT validation:* three independent patient populations with visual and quantitative HRCT and 24-month PFS data were combined and used for validation including: (1) a single center fHP research cohort (n= 120) seen at National Jewish Health (NJH) and evaluated in the ILD program between January 2018 and June 2025, (2) a single-center, prospective, randomized, pirfenidone clinical trial conducted at NJH in 40 patients with fHP (NCT02958917)^19^ conducted from June 2017 to April 2020 and (3) the Pulmonary Fibrosis Foundation Patient Registry (PFF-PR, n=56 with fHP),^20^ **Figure 1**.

*Change from baseline and 12-month HRCT validation:* Included only (1) the single-center research cohort (n= 120), and (2) the single-center trial (n= 40)^19^ since they had both baseline and 12-month HRCT and subsequent 12-month PFS data.

In all cohorts, axial, inspiratory, noncontrast chest HRCT scans, obtained within 60 days of the baseline and at 12 months, were selected for analysis. Scans with slice thickness >1.5 mm, axial slice spacing >0.0 mm, or incomplete representation of the lungs were considered inadequate and were excluded.

### Visual HRCT Scan Assessments

All HRCT scans from the primary and validation cohorts were independently reviewed by one experienced thoracic radiologist (D.A.L.) who was blinded to individual’s clinical status, demographic data, and HRCT date. All scans were classified according to three morphologic categories as outlined by the HP guidelines (typical, compatible or indeterminate for fHP).^5^ Scans were also classified as UIP-like (UIP or probable UIP) or non-UIP (indeterminate or other) according to the current American Thoracic Society and Fleischner Society guidelines on the diagnosis of IPF.^21,22^

### Quantitative CT Analysis and MIL

We utilized the DTA on technically adequate CT scans within each cohort to measure the overall extent of lung fibrosis (DTA fibrosis score),^23,24^ including reticular abnormalities, honeycombing, and ground-glass opacities, and the MIL-UIP expressed as a continuous score between 0.0 and 1.0, with larger values suggesting higher likelihood of UIP.

### Statistical Analysis

Descriptive statistics for baseline and longitudinal data were calculated. Continuous variables are reported as means with standard deviations, while categorical variables are expressed as counts with percentages.

Linear mixed-effects models were fit to assess how DTA overall fibrosis scores were associated with FVC and DLCO. For both parameters, separate models were fitted using the percentage predicted as the repeated measures outcome. We used the method of Neuhaus et al.^25^ to partition the association between DTA and each outcome into between- and within-subject components. This was done by transforming each individual’s repeated DTA measures into a person-level mean and a deviation from this mean at each of that patient’s observations; the regression coefficients for these two covariates capture the between- and within-subject components, respectively. Also included in each model was a random intercept for each participant to account for the correlation between repeated observations. Inference is based on t-tests using the Satterthwaite method for calculating degrees of freedom.

Kaplan–Meier plots were used to visualize PFS by groups defined by quartiles of DTA fibrosis score. Multivariable Cox analysis adjusted for age, sex, antigen exposure history (except in the PFF-PR cohort), smoking history, anti-fibrotic therapy and baseline FVC% was performed to assess the association between baseline variables and risk of 24-month PFS and the association between 12-month change in DTA, DTA at 12 months and PFS.

PFS was defined as the time from baseline HRCT to a decline in FVC of ≥10%, a decline in DLCO of ≥10%, lung transplantation or death. Analyses in the validation cohort mirrored those described above.

Statistical significance was set at α = 0.05. All analyses were performed using SAS version 9.4 and R version 4.4.2.

## RESULTS

### Study cohorts

Of the 137 eligible patients with fHP in the primary cohort, 134 were included in the baseline analyses and the analysis of the 12-month change in DTA (**Figure 1**). A total of 216 similarly defined individuals were included in the validation cohort (**Figure 1**). Demographic, clinical and HRCT characteristics for the primary and validation fHP cohorts are presented in **Table 1**.

**Table 1.** Baseline Characteristics of Primary and Validation Cohorts.

| Characteristic | Primary Cohort<br>(N=134) | Validation Cohort |  |  |  |
| --- | --- | --- | --- | --- | --- |
|  |  | Overall<br>(N=216) | NJH-HP<br>(n=120) | RCT-HP<br>(n=40) | PFF-PR<br>(n=56) |
| Age, years, mean (SD) | 69.4 (8.7) | 66.5 (8.6) | 66.7 (9.1) | 67.1 (5.7) | 66.3 (9.3) |
| Female, n (%) | 67(50.0) | 130 (60.2) | 77 (64.2) | 23 (57.0) | 30 (53.6) |
| Ever smoker, n (%) | 58 (43.2) | 89 (41.2) | 43 (35.8) | 21 (52.5) | 25 (44.6) |
| Identifiable antigen exposure, n (%) | 77 (57.5) | 85 (53.1) * | 52 (43.3) | 33 (82.5) | - |
| FVC% predicted | 72.5 (19.9) | 67.7 (17.6) | 71.9 (18.2) | 61.2 (11.8) | 65.0 (17.2) |
| DLCO% predicted | 59.9 (18.3) | 53.8 (23.1) | 61.4 (29.3) | 52.6 (14.9) | 45.0 (14.3) |
| Antifibrotic treatment | 51 (38.1) | 43 (19.9) | 11 (9.2) | 27 (67.5) | 5 (8.9) |
| HRCT UIP-like | 18 (13.4) | 33 (15.8) | 24 (20.0) | 0 | 9 (16.1) |
| HRCT HP pattern confidence, n (%) |  |  |  |  |  |
| Typical | 33 (24.6) | 123 (56.9) | 84 (70.0) | 20 (50.0) | 19 (33.9) |
| Compatible | 54 (40.3) | 60 (27.8) | 12 (10.0) | 20 (50.0) | 28 (50.0) |
| Indeterminate | 47 (35.1) | 33 (15.3) | 24 (20.0) | 0 | 9 (16.1) |
| DTA total fibrosis extent, mean (SD) | 28.3 (18.9) | 23.4 (17.6) | 20.15 (17.4) | 30.9 (14.2) | 24.8 (18.7) |
| DTA ground glass extent | 5.2 (2.5) | 5.8 (2.6) | 4.8 (1.8) | 89.0 (2.6) | 5.6 (2.2) |
| DTA honeycombing extent | 1.2 (3.5) | 1.2 (3.7) | 1.2 (3.9) | 1.8 (5.1) | 1.1 (2.3) |
| DTA reticulation extent | 26.5 (18.5) | 23.1 (17.5) | 20.4 (18.0) | 28.2 (13.7) | 23.8 (17.8) |
| MIL-UIP score (continuous), mean (SD) | 0.6 (0.3) | 0.6 (0.5) | 0.6 (0.6) | 0.7 (0.2) | 0.5 (0.30) |
| MIL-UIP score (binary), n (%) |  |  |  |  |  |
| ≤ 0.5 | 51 (38.1) | 135 (62.5) | 70 (58.3) | 33 (82.5) | 32 (57.1) |
| > 0.5 | 83 (61.9) | 81 (37.5) | 50 (41.7) | 7 (17.5) | 24 (42.9) |

\*92/160 (57.5%). Data not available for PFF-PR.
Abbreviations: NJH = National Jewish Health; RCT = randomized clinical trial; PFF-PR = Pulmonary Fibrosis Foundation Patient Registry.

The primary cohort had a mean age of 69.4 years (± 8.7), with an equal sex distribution. 43.1% were ever smoker and 57.5% had an identifiable inciting antigen exposure. The mean FVC was 72.5% (± 19.8), and the mean DLCO was 59.9% (± 18.3). CT appearances were characterized as typical pattern in 24.6% of cases, a compatible pattern in 40.3%, and an indeterminate pattern in 35.1%. The mean baseline DTA fibrosis score was 28.0 (± 18.8) and 61.9% had a MIL-UIP score >0.5.

The validation cohort had a mean age of 66.5 years (± 8.6), and 60% were female sex. 40% were ever smoker and 53.1% had an identifiable inciting antigen exposure in the combined NJH-HP and the RCT cohort. The mean FVC was 67.7% (± 17.6), and the mean DLCO was 53.8% (± 23.1). CT appearances chests were characterized as typical pattern in 55.9% of cases, a compatible pattern in 28.8%, and an indeterminate pattern in 15.3%. The mean baseline DTA fibrosis score was 23.8 (± 17.8) and 40% had a MIL-UIP >0.5.

In the primary and validation cohorts, patients with a MIL-UIP > 0.5 had higher DTA fibrosis scores compared to those with a MIL-UIP of ≤ 0.5 (35.9 ± 16.3 vs. 11.6 ± 12.0, *P* < 0.001 and 31.4 ± 16.5 vs. 10.0 ± 10.2, *P* < 0.001, respectively). Similarly, although the primary cohort had a small number of patients with a CT UIP-like compared to the validation cohort, in both cohorts, patients with a visual CT UIP-like pattern trended toward higher DTA fibrosis scores compared to those with a non-UIP-like pattern (35.6 ± 14.7 vs. 26.8 ± 19.2, *P =* 0.08 and 35.6 ± 20.8 vs. 21.6 ± 16.3, *P* < 0.001, respectively).

### Association between baseline visual CT HP pattern, MIL-UIP, baseline DTA and 24-Month PFS

In the primary and validation cohorts, baseline visual HRCT patterns of HP or UIP-like were not associated with 24-month PFS (**Table 2**). In contrast, the MIL-UIP classification was significantly associated with 24-month PFS when analyzed either continuously or dichotomously (**Table 3**). However, the MIL-UIP classifier did not significantly predict 24-month PFS after adjusting for DTA fibrosis score.

**Table 2.** HRs (95% CIs) for 24-months Progression-Free Survival, by Baseline Visual CT Pattern for the Primary and Validation Cohorts.

| Covariate | Primary Cohort N=134 |  |  | Validation Cohort N=216 |  |  |
| --- | --- | --- | --- | --- | --- | --- |
|  | HR | (95% CI) | P value | HR | 95% CI | P value |
| Age | 0.995 | (0.965, 1.026) | 0.762 | 1.004 | (0.981, 1.028) | 0.737 |
| Male | 1.058 | (0.627, 1.785) | 0.834 | 0.795 | (0.539, 1.172) | 0.246 |
| Ever smoker | 1.232 | (0.725, 2.090) | 0.441 | 1.114 | (0.757, 1.639) | 0.584 |
| Identifiable antigen exposure | 0.458 | (0.261, 0.804) | 0.007 | 0.636* | (0.406, 0.994) | 0.047 |
| FVC% predicted | 0.966 | (0.950, 0.983) | <0.001 | 0.981 | (0.969, 0.993) | 0.002 |
| DLCO% predicted | 0.965 | (0.948, 0.983) | <0.001 | 0.988 | (0.977, 1.000) | 0.051 |
| Antifibrotic treatment | 0.536 | (0.192, 1.497) | 0.234 | 2.710 | (1.051, 6.990) | 0.039 |
| HRCT pattern |  |  |  |  |  |  |
| UIP-like vs. non-UIP like | 0.805 | (0.324, 1.999) | 0.640 | 1.026 | (0.598, 1.762) | 0.926 |
| HP Pattern |  |  |  |  |  |  |
| Indeterminate vs. Compatible | 0.696 | (0.355, 1.366) | 0.292 | 1.019 | (0.615, 1.689) | 0.941 |
| Typical vs. Compatible | 0.781 | (0.360, 1.694) | 0.531 | 1.378 | (0.889, 2.136) | 0.151 |
| Typical vs. Indeterminate | 1.122 | (0.543, 2.318) | 0.756 | 1.352 | (0.769, 2.378) | 0.295 |

\*N = 160.

**Table 3.**
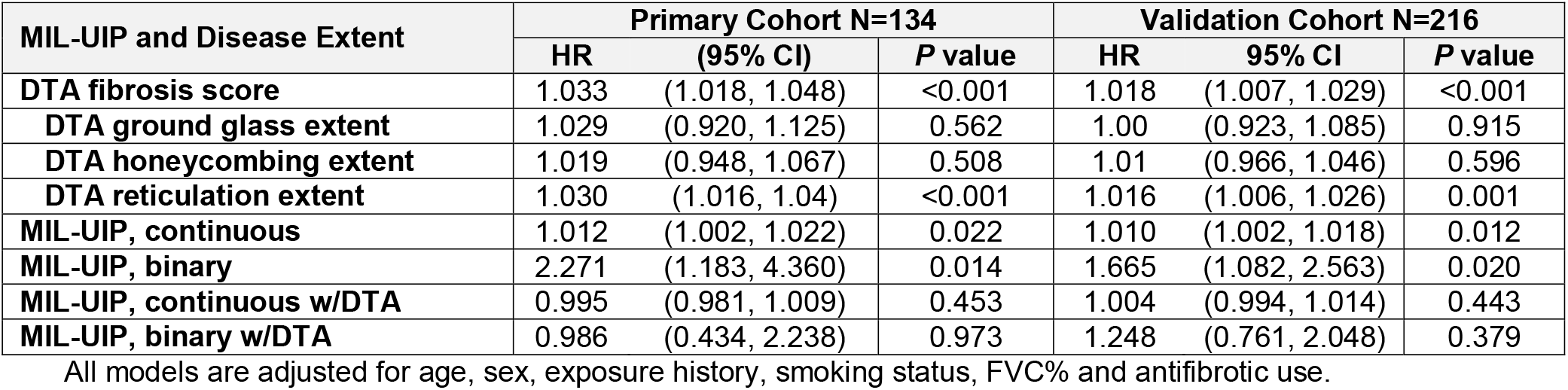
HRs (95% CIs) for 24-months Progression-Free Survival, by MIL-UIP and DTA for the Primary and Validation Cohorts.

DTA fibrosis scores at baseline were associated with decreased 24-month PFS in the primary (HR=1.03, 95% CI 1.018, 1.048, *P* < 0.001) and validation cohorts (HR=1.018, 95% CI 1.007, 1.029, *P* <0.001). Patients in the third or fourth DTA quartile had worse 24-month PFS compared to those in the lowest or first DTA quartile. (**Figure 2**).

**Figure 2.**
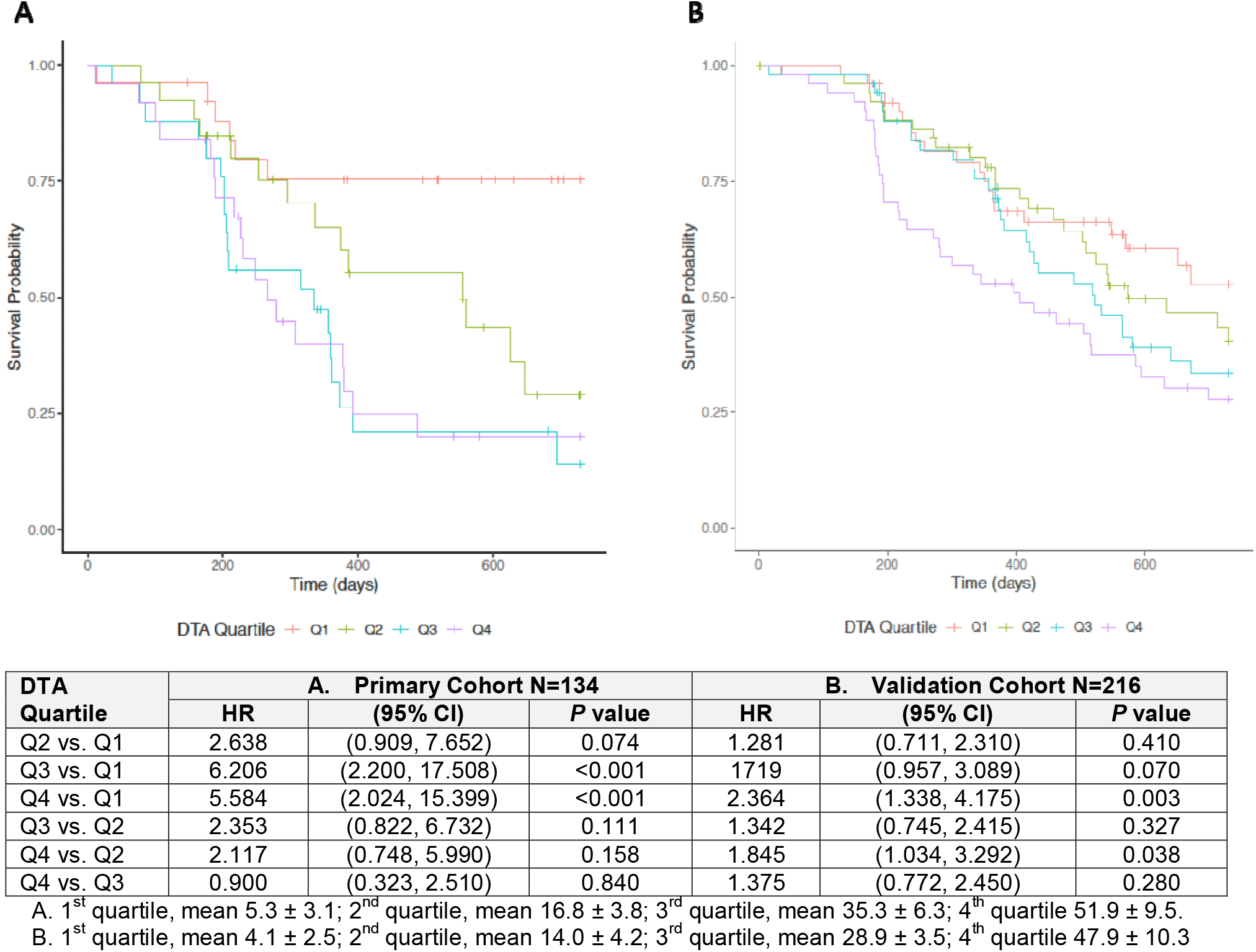
Kaplan–Meier plots showing progression-free survival by quartiles of baseline data-driven texture analysis fibrosis score at baseline computed tomography for the primary (A) and validation (B) cohorts.

### Association between DTA and PFTs at baseline and at 12-Month

In the primary cohort, the linear mixed model showed that differences in DTA fibrosis scores between individuals were closely associated with variations in FVC% at baseline and at 12 months. A 5-point increase in DTA fibrosis score across participants was associated with a 3.5 percentage point decline in FVC% (*P* <0.001) and a 3.1-point decline in DLCO% (*P* <0.001). Changes in DTA fibrosis score within an individual were also significantly associated with FVC% decline, with a 5-point increase in DTA fibrosis score within an individual resulting in a 1.8 percentage point decline in FVC (*P* <0.001) and a 2.5 percentage point decline in DLCO%. Similar results were observed in the validation cohort (**Table 4**).

**Table 4.** Linear mixed model results of the associations between DTA and both FVC% and DLCO% partitioned into between-subject (cross-sectional) and within-subject (longitudinal) analyses.

| Covariate | Between Subject Component |  |  | Within Subject Component |  |  |
| --- | --- | --- | --- | --- | --- | --- |
|  | Coef. | (95% CI) | P value | Coef. | (95% CI) | P value |
| **Primary Cohort (N=134)** |  |  |  |  |  |  |
| **FVC% predicted** | -0.709 | (-0.862, -0.556) | <0.001 | -0.358 | (-0.539, -0.178) | <0.001 |
| **DLCO % predicted** | -0.617 | (-0.749, -0.485) | <0.001 | -0.507 | (-0.751, -0.264) | <0.001 |
| **Validation Cohort (N=216)** |  |  |  |  |  |  |
| **FVC% predicted** | -0.561 | (-0.687, -0.436) | <0.001 | -0.582 | (-0.732, -0.431) | <0.001 |
| **DLCO % predicted** | -0.923 | (-1.078, -0.767) | <0.001 | -1.11 | (-1.491, -0.730) | <0.001 |

### Association between 12-Month Change in DTA, DTA at 12-Month and Subsequent PFS

In the primary cohort, an increase in DTA from baseline to 12 months was associated with shorter PFS in the following 12 months. A similar, though not statistically significant, trend in shorter PFS was observed in the validation cohort. In both the primary and validation cohorts, regardless of the baseline DTA, an increase in the DTA fibrosis score at 12 months was predictive of shorter PFS over the subsequent 12 months (**Table 5**).

**Table 5.** HRs (95% CIs) for 12-Month Change in DTA, DTA at 12-Month, and Subsequent 12-Month PFS.

| Covariate | Primary Cohort N=134 |  |  | Validation Cohort N=160 |  |  |
| --- | --- | --- | --- | --- | --- | --- |
|  | HR | (95% CI) | P value | HR | 95% CI | P value |
| **12 Month Change in DTA** | 1.067 | (1.011, 1.126) | 0.018 | 1.048 | (0.996, 1.103) | 0.071 |
| **DTA at 12 Months** | 1.029 | (1.005, 1.052) | 0.015 | 1.017 | (1.002, 1.031) | 0.022 |

All models are adjusted for age, sex, exposure history, smoking status, FVC% and antifibrotic use.

## DISCUSSION

We evaluated the prognostic performance of (1) baseline visual CT patterns of HP and UIP, as well as the MIL-UIP classification, and (2) DTA, both at baseline and over a 12-month period, and at 12 months in multicenter independent patient cohorts with fHP.

In contrast to other studies that did not account the presence of radiologic HP pattern or the seemingly more prognostically predictive UIP-like pattern in relation to the extent of fibrosis, a key finding in this study is that the visual HP or UIP pattern, or even the automated MIL-UIP score (greater than 0.5), does not provide additional prognostic information beyond the extent of lung fibrosis on HRCT by DTA in fHP. We demonstrate that quantitative scores of fibrosis extent hold greater prognostic significance than the identification of a diagnostically relevant visual CT fHP or UIP-like pattern. This suggests that assessing the extent of disease should enable a more appropriate stratification of fHP patients compared to relying solely on CT phenotypes. This approach may have significant implications for classifying fHP patients at risk of disease progression and for comparing treatment responses among fHP patients, as well as potentially other non-IPF ILDs, in clinical trials. Notably, to date, non-IPF phase 3 clinical trials have not incorporated the quantitative extent of disease as a secondary endpoint.

Our findings also indicate that among patients with fHP, the risk associated with radiologic UIP is related to the greater extent of lung fibrosis, which often corresponds with a more confident visual diagnosis of UIP. Indeed, our analysis revealed that the baseline classification utilizing the MIL-UIP algorithm^17^ was associated with higher DTA scores.

In several prior studies, as is commonly done in clinical practice, clinicians assess the presence and distribution of CT features, which inform the overall CT pattern and potentially future disease behavior. However, our study builds on prior quantitative CT studies evaluating a broader spectrum of ILDs^1,12,26-28^ and suggests that, in HP, a quantitative composite score may be superior to any of its individual components (i.e., honeycombing, reticulation, and ground glass) in predicting outcomes. This is likely because, compared to individual CT features of lung injury, the overall extent of disease serves as a better direct measure of how much of the lung’s functional and structural network has been compromised, indicating how close the lung is to its functional percolation threshold,^29^ a point that is more directly correlated with clinical decline than the visual appearance of CT features or pattern.

While the baseline DTA fibrosis score and other quantitative CT methods have been shown to be predictive of poor outcomes, including lung function decline and shorter survival in IPF,^15,16^ systemic autoimmune disease-associated ILD^14^ and overall mixed ILD populations,^13^ a novel finding in this study is that DTA change from baseline to 12-month as well as the DTA fibrosis score at 12 months, independent of baseline DTA scores, was associated with subsequent 12-month PFS in HP. This finding, similar to recent findings in subjects with a broader spectrum of fibrotic lung diseases,^13^ carries important implications, highlighting that access to a reproducible and visually unbiased predictor of 12-month disease outcome may enable clinicians to engage in informed discussions regarding prognosis with their patients earlier in the disease course, potentially leading to improved disease management and prognosis.

The strengths of this study include its prospective multicenter cohort design with a protocol-driven multidisciplinary HP diagnosis to ensure consistency across sites. The study’s aim of assessing DTA both at baseline and over time was pre-specified before data collection,^18^ thereby minimizing bias in hypothesis testing after the results were known. Also, our findings were replicated using a large, independent validation cohort supporting the external validity of our findings.

However, the generalizability of the study results is subject to some limitations. While the replication of the findings is encouraging, the HRCT scans were obtained as part of standard clinical care and lacked standardization in both the validation sample of the single-center study and the PFF-PR registry. Variability in CT reconstruction protocols, discrepancies in CT acquisition timing, and the presence of missing PFS data contributed to a reduction in sample size. This limitation may have adversely affected the validation of 12 month change in DTA and subsequent PFS by potentially introducing unmeasured confounding and selection biases. Also, the HRCT pattern was determined by a single radiologist. Visual assessment of the CT pattern can be subject to inter-observer variability, which may explain why we found that visual CT pattern evaluation may be less sensitive as a prognostic marker in patients with HP-UIP compared to the probability of UIP determined through the MIL-UIP. However, it is encouraging that both the visual UIP-like pattern and the results of the MIL-UIP classifiers were consistently replicated (**Table 3**).

## Conclusions

We demonstrated, in a multicenter study with independent fHP populations, that the baseline DTA fibrosis score, along with its change over 12 months and DTA at 12 months, are associated with subsequent 24-month and 12-month decreases in PFS, respectively, underscoring the importance of closely monitoring the extent of CT fibrosis over time to better forecast outcomes in patients with fHP. We also found that quantitative CT assessment of lung fibrosis extent is more prognostic than a UIP classifier or visual CT patterns of HP in patients with HP. This adds to the growing body of evidence suggesting that incorporating quantitative CT methods may enhance prognostic risk stratification in clinical practice and inform personalized patient care for patients with fHP.

## Data Availability

All data produced in the present work are contained in the manuscript.

